# Predicting self-harm at one year in female prisoners: a retrospective cohort study using machine learning

**DOI:** 10.1101/2023.09.20.23295770

**Authors:** Paul A Tiffin, Sant Leelamanthep, Lewis W Paton, Amanda E. Perry

## Abstract

**Background:** Self-harm and suicide are relatively overrepresented in incarcerated populations, especially in female prisons. Identifying those most at risk of significant self-harm could provide opportunities for effective, targeted interventions.

**Aims:** To develop and validate a machine learning-based algorithm capable of achieving a clinically useful level of accuracy when predicting the risk of self-harm in female prisoners.

**Method:** Data were available on 31 variables for 286 female prisoners from a single UK-based prison. This included sociodemographic factors, nature of the index offence, and responses to several psychometric assessment tools used at baseline. At 12-month follow-up any self-harm incidents were reported. A machine learning algorithm (CatBoost) to predict self-harm at one-year was developed and tested. To quantify uncertainty about the accuracy of the algorithm, the model building and evaluation process was repeated 2000 times and the distribution of results summarised.

**Results:** The mean Area Under the Curve (AUC) for the model on unseen (validation) data was 0.92 (SD 0.04). Sensitivity was 0.83 (SD 0.07), specificity 0.94 (SD 0.03), positive predictive value 0.78 (SD 0.08) and the negative predictive value 0.95 (0.02). If the algorithm was used in this population, for every 100 women screened, this would equate to approximately 17 ‘true positives’ and five ‘false positives’.

**Conclusions:** The accuracy of the algorithm was superior to those previously reported for predicting future self-harm in general and prison populations and likely to provide clinically useful levels of prediction. Research is needed to evaluate the feasibility of implementing this approach in a prison setting.

## Introduction

Self-harm includes various behaviours or actions of injuring or hurting oneself, either with or without suicidal intentions. Self-harm is strongly associated with the risk of completed suicide and death by misadventure (Chen, Tan et al. 2011). It also negatively impacts on social relationships and other aspects of life (Wu, Chang et al. 2013). Mental health issues significantly predispose individuals to self-harm and suicide (Singhal, Ross et al. 2014). Certain sociodemographic factors are also associated with an increased risk of self-harm and suicidality. These include socioeconomic disadvantage, female gender, childhood abuse, as well as other early life adverse experiences (Russell, Heron et al. 2019).

Self-harm has a prevalence of 6.4% in England, with a trend towards increased rates over time (McManus, Gunnell et al. 2019). The reported rates are substantially higher in imprisoned, compared to general populations. This is especially so for women prisoners with a reported prevalence of self-harm around 20-24% (Hawton, Linsell et al. 2014). There is evidence that prediction of self-harm is generally challenging with many approaches little better than chance, and not clinically useful (Franklin, Ribeiro et al. 2017). Moreover, mental health screening programmes of any sort are often hindered by relatively high number of ‘false positive’ cases that needlessly absorb resources and potentially increase anxiety in patients and clinicians (Paton and Tiffin 2022). However, the number of ‘false positives’ is related to the negative and positive predictive value of a test. This varies with the prevalence of the target condition. The high rates of self-harm in female prisoners thus provides an opportunity to develop a risk assessment system with potentially clinically useful levels of accuracy. Several interventions are available that could potentially be used to reduce the risk of self-harm and suicidality in individuals identified as being at especially high risk (Perry, Waterman et al. 2019, Winicov 2019).

Amongst prisoners the strongest predictors of self-harm in prisons have been reported to include; current or recent, or lifetime suicidal ideation, previous self-harm and any current psychiatric diagnosis (particularly depression and borderline personality disorder (Favril, Yu, Hawton, Fazel, 2020). The prison-specific environmental risk factors for self-harm include solitary confinement, disciplinary infractions and experiencing sexual or physical victimisation during imprisonment. Sociodemographic and criminological factors (such as nature of the index offence) have been reported to be only modestly associated with the risk self-harm in prison (Favril et al 2020).

There have been several prior attempts to develop screening and prediction approaches to identify those prisoners most at risk for self-harm. Previous studies do not always cite all the predictive accuracy metrics of interest (e.g., the positive predictive value). However, they do tend to report the Area Under the Curve (AUC) values for tools evaluated as potential screening instruments. In this context an AUC is generated from a Receiver Operator Characteristic (ROC) curve which plots the proportion of ‘false positives’ against ‘false negatives’ (i.e., sensitivity vs 1-specificity) for any given hypothetical threshold score used as a screening cut-off. As such, the AUC is an indicator of the test to act as an effective screening tool. That is, the AUC indicates whether there is likely to be a potential cut-score that offers an optimum, and possibly acceptable, trade-off between sensitivity and specificity. Formally, AUC values close to 1.00 indicating almost 100% accuracy whilst those around 0.5 would indicate a screening test performance that was no better than chance.

Though not a prison population as such; a previous small cohort study of patients (N=34) with previous criminal offending, drawn from a UK-based medium-secure mental health unit evaluated the ability of several instruments to predict violence and self-harm (Gray, Hill et al. 2003). These tools consisted of the Historical, Clinical, and Risk Management Scales (HCR-20), Psychopathy Checklist—Revised (PCL–R), Beck Hopelessness Scale (BHS) and the Brief Psychiatric Rating Scale (BPRS). Of these, only the BHS score statistically significantly predicted the occurrence of self-harm at 3-month follow-up, with an AUC of 0.86 cited. A larger scale study, of female UK prisoners (N=394) evaluated the ability of several instruments to predict self-harm at 4-year follow-up (Perry and Gilbody 2009). The tools used were the Beck Depression Inventory (BDI), the Beck Hopelessness Scale (BHS), and a novel instrument, known as the ‘Suicide Concerns for Offenders in Prison Environment’ (SCOPE). In terms of predicting future self-harm, at follow-up, the respective AUC values cited were 0.75 for the BDI, 0.59 for the BHS and 0.69 for the SCOPE tool.

A separate prospective cohort study, involving both male and female prisoners, evaluated the abilities of the Prison Screening Questionnaire (PriSnQuest), a modified Borderline Symptom List-23 (BSL-23), Self-Harm Inventory (SHI), Patient Health Questionnaire-9 (PHQ-9) and Clinical Outcomes in Routine Evaluation – Outcome Measure (CORE-OM) to predict self-reported self-harm at 6 month follow-up (Horton, Dyer et al. 2018). Those completing the questionnaires were already deemed to be at an increased risk of self-harm at recruitment.

Individually, each instrument had been established as having relatively poor ability to predict self-harm at follow-up in this cohort (Horton, Wright et al. 2014). Therefore, statistical analyses were conducted to develop two algorithms (for male and females separately) that only included scores from the most predictive items in the battery of psychometric instruments used. The resulting, novel scale scores had AUC values cited as 0.81 for males and 0.87 for female prisoners. However, the algorithms were developed and validated in the same dataset. Consequently, the AUCs cited will be somewhat inflated. Similarly, a separate study of 542 UK prisoners developed and tested a self-harm screening tool containing 11 items, with separate versions for men and women (Ryland, Gould et al. 2020). The score from the tool was reported to have an AUC of 0.67 for predicting self-harm in the 6-month follow-up period. The instrument was further modified by dropping two items and weighting the risk factors, based on the findings from a multivariable model. This resulted in an observed AUC of 0.84.

These studies demonstrate that it may be possible to achieve AUC values that are relatively high in the context of mental health screening programmes in the secure estate. However, they generally share some key weaknesses, in that they fail to communicate how such AUCs translate into actual raw numbers of true and false positives, given the prevalence of the target condition. Secondly, such studies tend to validate their tools the same data that has been used to develop them.

Machine learning is a subfield of artificial intelligence (AI) that can offer a flexible approach to prediction. Complex non-linear relationships and interactions in data can be captured more easily by machine learning than classical statistical approaches. Machine learning also capitalises on advances in computing power so that very large numbers of models can be built, tuned, combined and evaluated in order to optimize the predictive accuracy of an algorithm. Machine learning has been applied to self-harm prediction to explore the possibility of improving accuracy. A review of machine learning for the prediction of self-harm, in general, reported that studies (in the community) usually reported moderate accuracy (AUC values of 0.74– 0.877)(Nordin, Zainol et al. 2022).

Machine learning has previously been applied to predicting self-harm in a population of 353 US-based prisoners (roughly 40% female) in substance misuse programmes (Horvath, Dras et al. 2021). Using a variety of algorithms the authors trained various models on cross-sectional data, attempting to predict self-harming behaviour. Including suicidal ideation resulted in an AUC of 0.955, though removing this predictor reduced this value to 0.875. The main weakness of this study was its cross-sectional nature.

Thus, the primary aim of the present study was to develop a machine learning based algorithm that was capable of accurately prospectively predicting self-harm over the medium term in female prisoners. A secondary aim was to identify the strongest predictors (‘features’) of self-harm in this group. The findings would be used to develop an approach that could then be evaluated for feasibility and clinical effectiveness in a woman’s prison.

## METHODS

### Data availability

Data were available for 286 women from a prison in the North of England (see Figure 1). Data were collected between January and April 2003 as part of a larger study of self-harm assessment in prisons (Perry and Olason 2009). The administration involved voluntary purposive cross samples of participants that were in prison on the day of administration. Administration was self-report and completed during prison ‘lock down’ over a series of successive lunchtime periods. A favourable ethical opinion for the original study was granted from the Ethical Committee at the University of York and a Governor at each prison.

**Figure 1.**
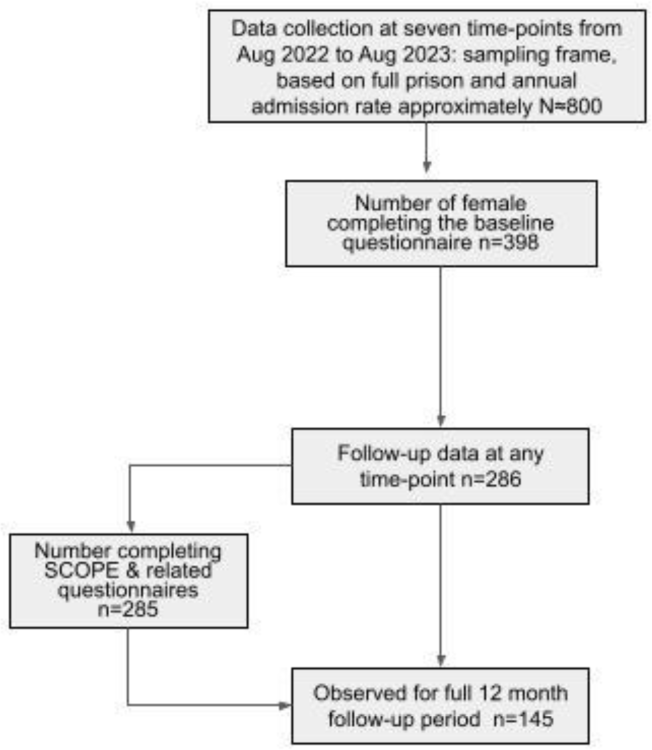
Flowchart of study participants.

Purposive samples (available on the day of interview) of female prisoners were approached to take part in the study. Eligible participants were identified as those at risk (i.e., had a previous history of either attempted suicide or nonfatal self-harm behaviour) between March 1 and July 31, 2002.

#### Outcome variable

Suicide or non-fatal self-harm was recorded at one year follow-up according to the prison records. Each participant was checked in the database of records including the Assessment Care in Custody and Teamwork (ACCT), National Offender Management Information System (NOMIS), and an ‘F2052SH form’ used to record incidence of self-harm. Recording of an incident in any one of these records was classed as an incident of self-harm. The period of ‘observation in prison’ was calculated from date of the administration of questionnaires to date of release or transfer. Therefore it was important to establish the length of time the participating women would have been observed in prison for, during which any self-harm would have been recorded.

#### Predictor variables

The predictor variables included in the model were baseline demographic characteristics (age in years, self-identified ethnicity), nature of the index offence, recorded history of self-harm intent and attempt, and summed scores from the validated psychometric assessment tools. These included: the Beck Depression Index (BDI), Beck Hopelessness Scale (BHS), and the Suicide Concerns for Offenders in Prison Environment (SCOPE) Tool. These are further described in the related studies (Perry and Gilbody 2009, Perry and Olason 2009). Missing values for predictor variables were imputed using a single hot deck imputation using the hot deck v1.2 package in R (Gill, Cranmer et al. 2022).

#### Machine learning approach

Our predictive models were built using gradient boosted decision trees via the CatBoost package v1.2 in R (Dorogush, Ershov and Gulin 2018). Gradient boosted decision tree models consist of an ensemble of decision trees, aiming to optimise model performance. This approach combines a number of methodological approaches to prediction; the use of decision trees; ‘ensembling’—where numerous slightly differing models are created, and the results averaged, and; ‘boosting’ where the algorithm successively focuses on the observations where the outcome is increasingly difficult to predict. By combining all three approaches, gradient boosted trees tend to outperform other algorithms for prediction in small and medium-sized data sets. In addition, CatBoost was specifically selected for this study as the software recodes categorical variables to numeric in a way which reflects their observed relationship with the outcome of interest. This can potentially increase the amount of information available to the predictive model. In our data there were several categorical variables, some of which were not *a priori* ordered, such as type of index offence. Thus, CatBoost offered a particular advantage in this context, often outperforming other machine learning approaches in structured (numeric) datasets (Hancock and Khoshgoftaar 2020).

The CatBoost algorithm has several key ‘hyperparameters’ that can be changed to improve predictive accuracy and generalisability: including the number of decision trees to grow, and; the number of variables to select at each node of the trees; the ‘learning rate’, and; the number of iterations of each learning round. The process of choosing hyperparameter settings is known as ‘tuning’. Ideally machine learning models should be tuned on a separate portion of data to reduce the risk of overfitting, which impedes generalisability. However, in this case, the dataset was relatively small and so to avoid the need for tuning a specific set of hyperparameters we built an ensemble of five CatBoost learners with a range of tuning settings. These settings were *a priori* selected based on the experience of using CatBoost for predictive tasks. For example, smaller learning rates tend to work better with more iterations. Thus, a spread of hyperparameters were used for five different CatBoost algorithms. The final prediction (self-harm at one year yes/no) was made by combining the outputs of the five algorithms via a majority vote.

When predicting relatively uncommon outcomes it is important to stop the algorithm focusing on achieving high accuracy by mainly predicting the most prevalent outcome (in this case, no self-harm at one year). For this reason, class weights were used to address the imbalance in outcomes (i.e., approximately 20% of the sample had self-harm reported at one year).

Data was randomly split, stratified by outcome, two-thirds of the data being used to train the algorithm and the remaining third used as the ‘hold-out’ (validation) dataset. Due to the stochastic nature of this algorithm development (e.g., dataset splitting, imputation, etc.) we repeated the entire process 2000 times, and the performance metrics were stored for each run. The overall performance of the models was evaluated by calculating the mean accuracy metrics over the 2000 iterations. This process is illustrated in Figure 2.

**Figure 2.**
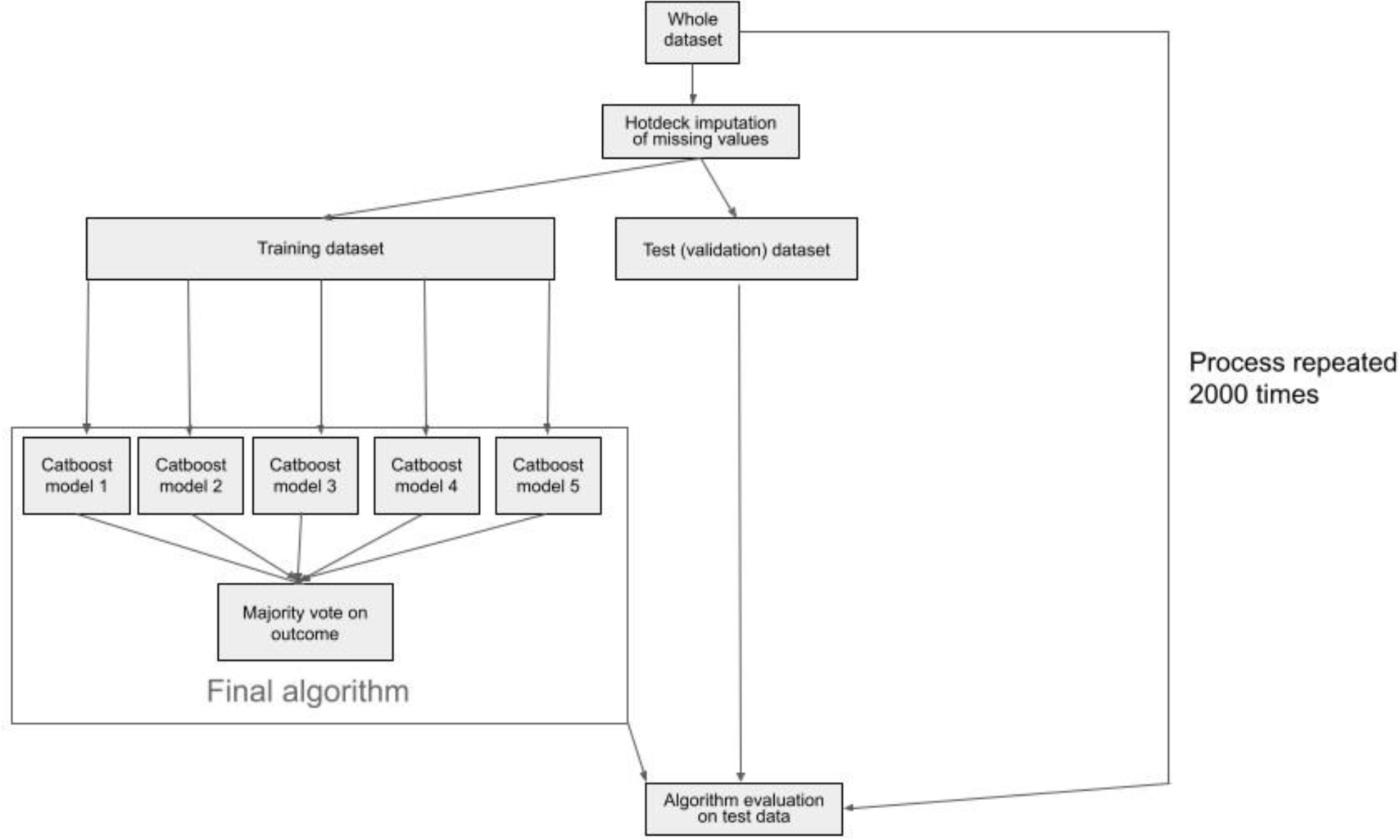
Flowchart of the machine learning model building and validation process.

Whilst around a quarter of the women in the sample had been observed in the prison for at least three months (the period) only 54% (155/286) had been so for six months or more. The minority of participants (39%, 112/286) had been observed for 12 months or more. The length of observation within a prison setting is a potential confounding variable in that it may mediate the relationship between the predictors and the probability of recording at least one act of self-harm. Unlike with survival analysis there was no direct way of controlling for the variable ‘exposure’ time in this machine learning context. For this reason, a sensitivity analysis was conducted, repeating the model building and evaluation using only data from the 145 women who either had self-harm reported in the first year of follow-up, or had been observed for at least a year in prison.

## Results

The demographic characteristics of the sample of female participants used for the analyses are shown in Table 1.

**Table 1.** Characteristics of the 286 participants in the study sample.

| Variable | Description | Missing |
| --- | --- | --- |
| Age | Mean 28.78 (SD 8.10)<br>years | n=1 |
| Ethnicity | 'White': 257 (91%)<br>'Black': 15 (5%)<br>'Mixed'/'Other': 10 (4%) | n=4 |
| Duration of follow-up in first year after baseline assessment | < 1 month: 26 (10%)<br>1 to 3 months: 44 (17%)<br>3 to 6 months: 36 (14%)<br>6m to 1 yr: 43 (16%)<br>≥1 year: 112 (43%) | n=25 |
| On remand? | Yes 167 (59%)<br>No 117 (41%) | n=2 |
| Index offence category | Assault 3 (1%)<br>Burglary 15 (5%)<br>Drug related 30 (10.5%)<br>Robbery 14 (5%)<br>Theft 30 (10.5%)<br>Other 194 (68%) | n=0 |

Model performance, averaged over 2000 iterations, in predicting self-harm within 12 months is shown in Table 2. As the distribution of values were approximately normally distributed (on inspection of quantile plots) the means and associated standard deviations (SDs) are provided. Note, as there was marked class imbalance, ‘balanced accuracy’ is shown, calculated as (sensitivity + specificity)/2.

**Table 2.**
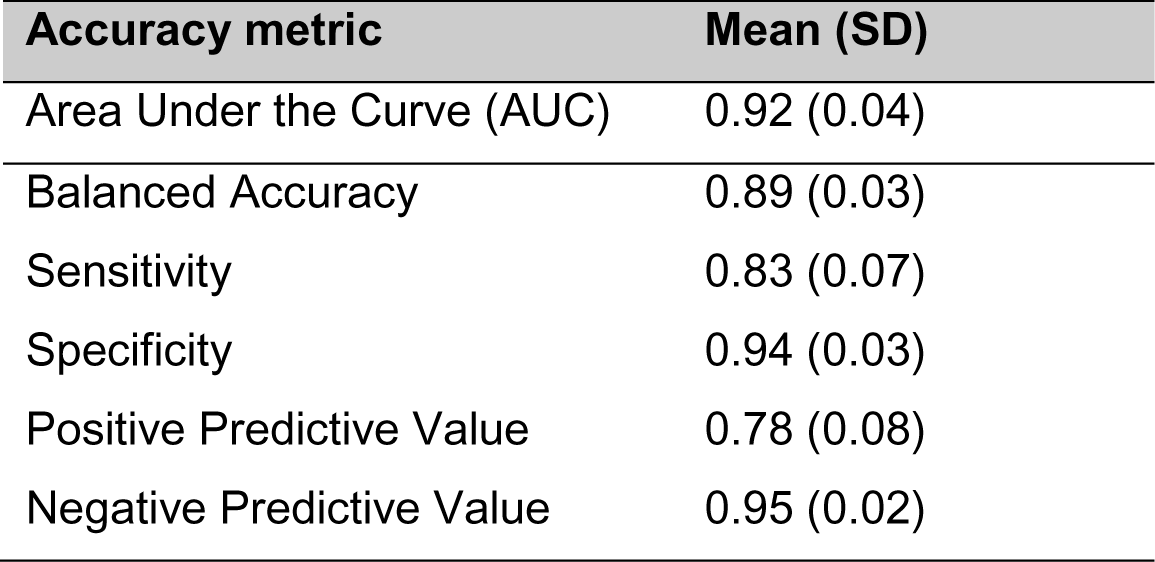
The average accuracy metrics for the performance of the algorithm on the validation (test) data.

| Accuracy metric | Mean (SD) |
| --- | --- |
| Area Under the Curve (AUC) | 0.92 (0.04) |
| Balanced Accuracy | 0.89 (0.03) |
| Sensitivity | 0.83 (0.07) |
| Specificity | 0.94 (0.03) |
| Positive Predictive Value | 0.78 (0.08) |
| Negative Predictive Value | 0.95 (0.02) |

The CatBoost algorithm does not produce interpretable models as such. However, the output for each run produced ‘importance’ metrics for the predictors. This is a normalised score for each variable which describes how much the prediction changes if the value of the predictor changes. In Table 3 we provide the mean importance scores for the predictors, averaged over the 2000 runs. As can be seen, the strongest predictor of self-harm among prisoners are any reported incidence of self-harm recorded on the prison system.

**Table 3.**
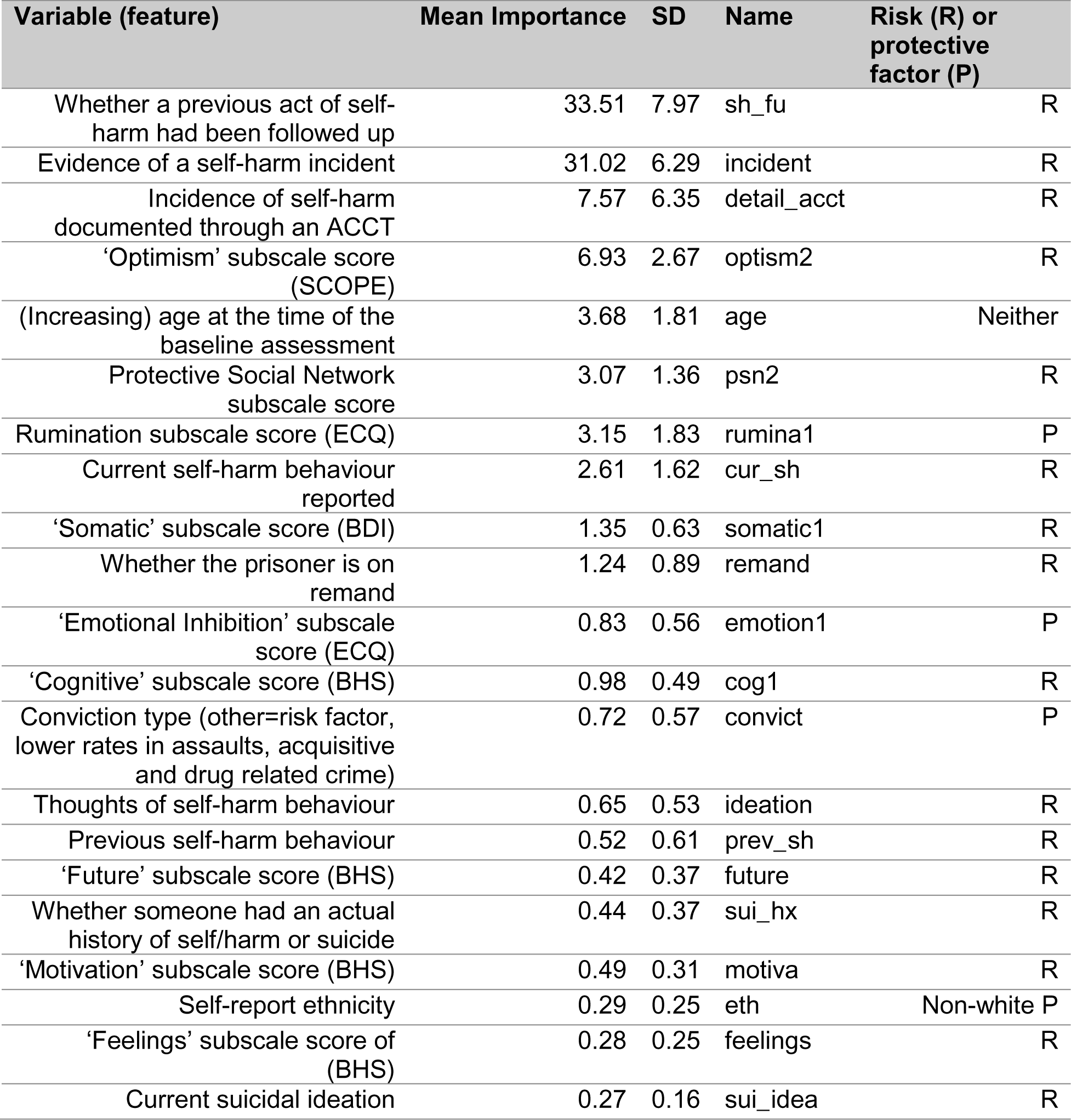
Mean importance of the predictor variables (features) in relation to the algorithm making predictions of future self-harm risk.

Assuming a prevalence of self-harm at one year of 20% in this population our results indicate that for every 100 women screened approximately 17 would be ‘true positives’ and five ‘false positives’. This level of precision may make such a process feasible in that only the minority of women with ‘false positive’ would need further assessment or to be offered interventions.

The results of the sensitivity analysis, using only data from the 145 women observed for at least one year in prison, or with self-harm reported, were very similar to those from the wider dataset. In this regard the mean AUC was modestly higher at 0.93 (compared to 0.92). As expected, due to the smaller numbers, the SD for the AUC was slightly larger than that derived from the wider dataset at 0.04 (compared to 0.03).

## Discussion

Using this machine learning approach, we were able to predict acts of self-harm at 12 month follow-up in this high-risk population with a relatively high degree of accuracy. Importantly the average positive predictive value observed for the validation dataset was relatively high at 79%.

Although not directly comparable, due to the different study samples, we noted the AUC values derived from our machine learning approach (at an average of 0.92) were considerably higher than that cited for the most predictive screening questionnaire (the SCOPE) in the earlier study, at 0.69 (Perry and Gilbody, 2009). No doubt this is because information from all the questionnaires, as well as demographic and criminal justice factors were used by the machine algorithm to make the predictions. The AUC we derived in our validation data is higher than the average reported for machine learning studies generally predicting self-harm (Nordin, Zainol et al. 2022). The AUC we observed for our algorithm also compares favourably with that of 0.84 cited by a previous study which developed a risk prediction model for self-harm in male prisoners (Ryland, Gould et al. 2020). However, in the latter case the authors did not estimate the performance of their algorithm in independent data, even using an internal resampling approach such as bootstrapping. Thus, the AUC cited in this case is likely to be an overestimate of the predictive ability of the model in a sample not used to develop and refine it. However, an earlier, small study involving 34 male in-patients in a medium secure unit also reported an AUC of 0.84 for the ability of the BHS scores to predict self-harm over a 3 month period (Gray, Hill et al. 2003). Thus, to date, our study reports the highest level of accuracy of any seeking to prospectively predict self-harm in a specific population.

According to the ‘importance metrics’, our algorithm relied most on information relating to previous suicidal ideation and self-harming behaviour when making the predictions. This is unsurprising, given that previous behaviour tends to be the most powerful predictor of future behaviour. There were however, some less expected predictors ranked in the top 10 importance metrics. These include whether an ACCT had been completed. This may have been a marker of the level concern from prison staff; although prison policy suggests that an ACCT document should be ‘opened’ regardless of severity. Also, age at baseline was ranked fifth but, on logistic regression did not seem to show a log linear relationship with risk of self-harm.

Thus, given that CatBoost is a tree-based algorithm the relationship with age and the primary outcome must have been non-linear and/or significantly interacted with other predictor variables. Increasing optimism, as rated by the SCOPE questionnaire, also seemed to be a risk factor for self-harm, according to the algorithm. It may have been that those who were more optimistic at baseline were more prone to acts of self-harm if subsequently disappointed or if any hopes were unrealised.

### Strengths and limitations

The main limitation of this study was the confidence relating to the ability to capture the outcome of interest. That is, the outcome of significant self-harm was recorded if it was self-reported by a participant and/or a member of prison staff. It therefore may have been affected by reporting bias. Moreover, this outcome was only recorded for those women who remained in prison at 12-month follow-up. This will inevitably have restricted the range of participants where an outcome could be observed, to those serving longer sentences. However, the results of our sensitivity analysis, which used only data from women who were observed for the full 12-month follow-up period were almost identical to those for the wider dataset.

We were able to validate our final algorithm using an ‘internal’ validation set, as well as quantify uncertainty over its performance by iterating the process many times. However, ideally, validation should have been carried out on a completely independent, external dataset, from the sample used to develop the algorithm. In addition, although we were able to quantify uncertainty the overall dataset was relatively small for a machine learning study. Thus, the 95% credible intervals derived from the iterative model building and testing process are relatively wide. Thus, a larger dataset would have provided further study power to estimate the likely actual precision of the predictive algorithm.

The machine learning approach we used would generally be considered ‘state of the art’. Boosted trees, and CatBoost in particular, takes advantage of categorical predictors. Thus it often outperforms other methods, including deep learning, in small to medium sized structured data sets (Grinsztajn, Oyallon and Varoquaux 2022). Moreover, we accommodated the unbalanced outcome (i.e., fewer individuals with self-harm present than absent in the dataset) by using class weights. This ensured that the algorithm did not mainly focus in the ‘easy’ task of predicting ‘non-self-harm’.

In contrast to most other published studies that employ machine learning, we have provided detailed and transparent descriptions of our methods. Indeed, in keeping with ‘Open Science’ principles we have made both our code and our analytic dataset publicly available. We have also attempted to anticipate the publication of the TRIPOD-AI consensus guidelines that will provide standards for the reporting of predictive and prognostic studies employing machine learning or artificial intelligence approaches (Collins, Dhiman et al. 2021).

### Implications for policy and practice

The data used to develop our predictive algorithm were derived from routinely available health, demographic and criminal justice data, as well as the scores derived from easily administered, fairly brief ‘paper and pencil’ questionnaires. The accuracy of our predictive algorithm, and specifically the positive predictive value, was superior to previously published self-harm prediction methods. This means that, in practice, where there is a relatively high prevalence of self-harm, the absolute numbers of ‘false positives’ will be relatively low. In the present case the ratio of true to false positive cases is at a level that may permit such an algorithm to support the care of women in criminal justice settings. However, our algorithm is relatively computationally intensive and its workings are not easily interpretable. The importance metrics provide information about which factors are particularly weighted within the component decision trees. However, the inner workings of the algorithm remain relatively opaque to humans. Thus, given the limited technological infrastructure typically found in prisons, it is not clear how easily implemented the algorithm would be. However, it may be that a simple set of decision tree rules, using four or five key variables, may work reasonably well, even if they lacked the level of accuracy of a more complex algorithm. Using a tree-based rule system would have the advantage of being easily interpretable, and, in theory, implemented using a paper and pencil flow-chart system. This may be more realistic in a prison setting.

Of course, if an individual is identified as at relatively high risk of self-harm there should be policies aimed at addressing this issue. This could involve identifying and treating co-existing conditions such as depression, anxiety and/or psychosis. The evidence for effective clinical interventions for self-harm in the absence of mental illness, or in the context of emotionally unstable (‘borderline’) personality disorder is somewhat patchy. However, there are some randomised controlled trials that report the effectiveness of psychological treatments including Dialectical Behavior Therapy, and Mentalisation-based Therapy (Stoffers, Völlm et al. 2012), though the former may be more effective than the latter (Barnicot and Crawford 2019). Thus, high-risk individuals could be targeted for individual or group therapy aimed at improving their emotional regulation and distress tolerance and reducing their reliance on self-harm as a coping strategy.

### Directions for future research

This study has demonstrated that the use of machine learning to capitalise on a wide range of routine and easily obtained information can achieve relatively high accuracy levels when predicting self-harm in a high-risk population. Our findings should be reproduced in an independent sample of female prisoners. Also, it may be that similar approaches will be fruitful in male prisoners, also at high risk of self-harm and suicide. Ultimately, however, what is required is a randomised, or quasi-experimental study, able to demonstrate that implementation of a screening/prediction interventions in a prison setting ultimately leads to better outcomes for prisoners, in terms of lower risks of self-harm, misadventure and suicide over the medium to long term. The screening itself is likely to be part of a complex intervention that will require organisational change and access to effective therapeutic interventions. Evaluating such an intervention will also require an economic component. However, given the high costs to the prison estate and associated health services of self-harm and suicidality, even interventions with modest effect-sizes are likely to be cost-effective.

### Conclusions

In this sample of female prisoners, a machine learning algorithm was able to predict, with potentially clinically useful accuracy, the risk of self-harm at 12-month follow-up. Further work should be carried out to implement such risk assessment systems in the prison system. However, translating such assessment processes into actual benefit to the individual prisoners, prison and wider society will be challenging. This will involve linking such assessment programmes to access to effective interventions.

## Data availability

The analytic dataset and code used in this analysis are available from https://github.com/pat512-star/prison_sh

## References

Barnicot, K. and M. Crawford (2019). “Dialectical behaviour therapy v. mentalisation-based therapy for borderline personality disorder.” Psychol Med 49(12): 2060–2068.

Chen, V. C. H., H. K. L. Tan, C.-Y. Chen, T. H. H. Chen, L.-R. Liao, C. T. C. Lee, M. Dewey, R. Stewart, M. Prince and A. T. A. Cheng (2011). “Mortality and suicide after self-harm: community cohort study in Taiwan.” The British Journal of Psychiatry 198(1): 31–36.

Collins, G. S., P. Dhiman, C. L. A. Navarro, J. Ma, L. Hooft, J. B. Reitsma, P. Logullo, A. L. Beam, L. Peng and B. Van Calster (2021). “Protocol for development of a reporting guideline (TRIPOD-AI) and risk of bias tool (PROBAST-AI) for diagnostic and prognostic prediction model studies based on artificial intelligence.” BMJ open 11(7): e048008.

Dorogush, A. V., V. Ershov and A. Gulin (2018). “CatBoost: gradient boosting with categorical features support.” arXiv preprint arXiv:1810.11363.

Franklin, J. C., J. D. Ribeiro, K. R. Fox, K. H. Bentley, E. M. Kleiman, X. Huang, K. M. Musacchio, A. C. Jaroszewski, B. P. Chang and M. K. Nock (2017). “Risk factors for suicidal thoughts and behaviors: A meta-analysis of 50 years of research.” Psychol Bull 143(2): 187–232.

Gill, J., S. Cranmer, N. Jackson, A. Murr, D. Armstrong and S. Heuberger (2022). hot.deck: Multiple Hot Deck Imputation.

Gray, N. S., C. Hill, A. McGleish, D. Timmons, M. J. MacCulloch and R. J. Snowden (2003). “Prediction of violence and self-harm in mentally disordered offenders: A prospective study of the efficacy of HCR-20, PCL-R, and psychiatric symptomatology.” Journal of Consulting and Clinical Psychology 71(3): 443–451.

Grinsztajn, L., E. Oyallon and G. Varoquaux (2022). “Why do tree-based models still outperform deep learning on typical tabular data?” Advances in Neural Information Processing Systems 35: 507–520.

Hancock, J. T. and T. M. Khoshgoftaar (2020). “CatBoost for big data: an interdisciplinary review.” Journal of big data 7(1): 1–45.

Hawton, K., L. Linsell, T. Adeniji, A. Sariaslan and S. Fazel (2014). “Self-harm in prisons in England and Wales: an epidemiological study of prevalence, risk factors, clustering, and subsequent suicide.” Lancet 383(9923): 1147–1154.

Horton, M., N. Wright, W. Dyer, A. Wright-Hughes, A. Farrin, Z. Mohammed, J. Smith, T. Heyes, S. Gilbody and A. Tennant (2014). “Assessing the risk of self-harm in an adult offender population: an incidence cohort study.” J Health Technology Assessment 18(64): 1–151, vii.

Horton, M. C., W. Dyer, A. Tennant and N. M. J. Wright (2018). “Assessing the predictability of self-harm in a high-risk adult prisoner population: a prospective cohort study.” Health Justice 6(1): 18.

Horvath, A., M. Dras, C. C. W. Lai and S. Boag (2021). “Predicting Suicidal Behavior Without Asking About Suicidal Ideation: Machine Learning and the Role of Borderline Personality Disorder Criteria.” Suicide and Life-Threatening Behavior 51(3): 455–466.

McManus, S., D. Gunnell, C. Cooper, P. E. Bebbington, L. M. Howard, T. Brugha, R. Jenkins, A. Hassiotis, S. Weich and L. Appleby (2019). “Prevalence of non-suicidal self-harm and service contact in England, 2000&#x2013;14: repeated cross-sectional surveys of the general population.” The Lancet Psychiatry 6(7): 573–581.

Nordin, N., Z. Zainol, M. H. Mohd Noor and L. F. Chan (2022). “Suicidal behaviour prediction models using machine learning techniques: A systematic review.” Artificial Intelligence in Medicine 132: 102395.

Paton, L. W. and P. A. Tiffin (2022). “Technology Matters: Machine learning approaches to personalised child and adolescent mental health care.” Child and Adolescent Mental Health 27(3): 307–308.

Perry, A., M. G. Waterman, A. House, A. Wright-Hughes, J. Greenhalgh, A. Farrin, G. Richardson, A. K. Hopton and N. Wright (2019). “Problem-solving training: assessing the feasibility and acceptability of delivering and evaluating a problem-solving training model for front-line prison staff and prisoners who self-harm.” BMJ Open 9(10): e026095.

Perry, A. E. and S. Gilbody (2009). “Detecting and predicting self-harm behaviour in prisoners: a prospective psychometric analysis of three instruments.” Soc Psychiatry Psychiatr Epidemiol 44(10): 853–861.

Perry, A. E. and D. T. Olason (2009). “A New Psychometric Instrument Assessing Vulnerability to Risk of Suicide and Self-Harm Behaviour in Offenders:Suicide Concerns for Offenders in Prison Environment (SCOPE).” International Journal of Offender Therapy and Comparative Criminology 53(4): 385–400.

Russell, A. E., J. Heron, D. Gunnell, T. Ford, G. Hemani, C. Joinson, P. Moran, C. Relton, M. Suderman and B. Mars (2019). “Pathways between early-life adversity and adolescent self-harm: the mediating role of inflammation in the Avon Longitudinal Study of Parents and Children.” 60(10): 1094–1103.

Ryland, H., C. Gould, T. McGeorge, K. Hawton and S. Fazel (2020). “Predicting self-harm in prisoners: Risk factors and a prognostic model in a cohort of 542 prison entrants.” Eur Psychiatry 63(1): e42.

Singhal, A., J. Ross, O. Seminog, K. Hawton and M. J. Goldacre (2014). “Risk of self-harm and suicide in people with specific psychiatric and physical disorders: comparisons between disorders using English national record linkage.” J R Soc Med 107(5): 194–204.

Stoffers, J. M., B. A. Völlm, G. Rücker, A. Timmer, N. Huband and K. Lieb (2012). “Psychological therapies for people with borderline personality disorder.” Cochrane Database Syst Rev 2012(8): Cd005652.

Winicov, N. (2019). “A systematic review of behavioral health interventions for suicidal and self-harming individuals in prisons and jails.” Heliyon 5(9): e02379.

Wu, C.-Y., C.-K. Chang, H.-C. Huang, S.-I. Liu and R. Stewart (2013). “The association between social relationships and self-harm: a case–control study in Taiwan.” BMC Psychiatry 13(1): 101.

